# Robust Performance of Culture, qPCR, and Genomic Approaches for Shigella Serotyping in a Pediatric Surveillance Cohort

**DOI:** 10.64898/2026.01.05.26343460

**Authors:** Francesca Schiaffino, Craig T. Parker, Lucero Romaina Cachique, Paul F. Garcia Bardales, Jie Liu, Pablo Peñataro Yori, Kerry K. Cooper, Ben Pascoe, Patricia Pavlinac, Eric Houpt, Maribel Paredes Olortegui, Margaret N. Kosek

**Author notes:** **Author Correspondence to**: Margaret N. Kosek; University of Virginia, Division of Infectious Diseases, International Health, and Public Health Sciences; Address: 345 Crispell Dr, Rm 2525, Charlottesville, VA.

## Abstract

**Background:** *Shigella* causes severe diarrheal disease, and *S. flexneri* and *S. sonnei* are the targets for multivalent vaccine development. Culture-based agglutination has been the gold standard for serotyping, but it is limited by logistics, subjectivity, and the availability of antisera for emerging serotypes. Newer methods, including a qPCR-based approach and whole-genome sequencing offer alternatives, but their performance in *Shigella* endemic populations are not well documented.

**Methods:** *Shigella* isolates obtained from the Enterics for Global Health (EFGH) study in Iquitos, Peru were simultaneously serotyped using four methods: culture-based agglutination, isolate-based qPCR serotyping, stool-based qPCR serotyping and WGS using the in-silico tool ShigaPass. The definitive adjudicated serotype was established by an expert analysis of the WGS data, involving the mapping of sequence reads to known O-antigen biosynthesis and modification genes to identify key mutations.

**Results:** Results from all four serotyping methods were available for 107/114 isolates. Accuracy for vaccine subtypes *S. flexneri* 1b, 2a, 3a, 6, and *S. sonnei*, ranged from 93.3-100% for all methods. Complete concordance between methods was noted in 83/107 isolates, while 24/107 (22.4%) exhibited at least one discrepancy. Most discrepancies derived from *S. flexneri* serotypes Y, Yv and 1a. Agglutination misclassified eight Y/Yv isolates as 4a, and six isolates correctly classified as 1a by agglutination were classified as 1b by the other methods, a discrepancy associated with a nonsense mutation in the *oac* gene.

**Conclusion:** All four serotyping methods achieved acceptable accuracy for *Shigella* vaccine efficacy evaluation. Although discrepancies are infrequent, WGS provides information of their genomic basis.

**Importance:** Shigella serotyping is critically important for the evaluation of future multivalent vaccines, of which there are several in advanced stages of development, as well as for monitoring of emerging Shigella serotypes. Culture based agglutination is the most widely used serotyping method, yet its successful implementation is associated with key logistical constraints. This study compares culture-based, qPCR-based, and whole-genome sequencing serotyping methods using isolates from a *Shigella*-endemic population in Peru. The study demonstrates that molecular and genomic approaches achieve high accuracy for vaccine-relevant serotypes and identifies the genomic basis of serotyping discrepancies. These methods would also reduce variation and improve data quality for future vaccine trials and epidemiologic surveillance. Ultimately, this work informs clinical microbiology laboratories and public health programs that seek a reliable and scalable alternative to traditional serotyping methods.

## Introduction

Diarrheal diseases continue to contribute to 500,000 deaths annually, globally (1). *Shigella* spp. is estimated to be the leading cause of death due to diarrhea as a result of a bacterial pathogen, and several vaccines are currently being developed for its control and prevention(2–4). The majority of cases of *Shigella* are caused by *Shigella flexneri* (comprised of 16 serotypes) and *S. sonnei* (comprised of a single serotype), and current strategies aim to develop multivalent vaccines with antigens restricted to these two serogroups (5).

Vaccine development and evaluation require robust and durable typing strategies for organisms such as *Shigella flexneri* that have multiple serotypes. The lipopolysaccharide (LPS) of *Shigella* consists of a relatively well conserved core lipid A structure, a species specific O-antigen backbone of polysaccharides encoded by the *rfb* genes, and a diverse set of terminal O-polysaccharide modifications that originate from lysogenic phages and plasmids (6–13). The terminal O-antigen modifications account for the antigenic specificity described by current typing strategies (14).

The culture-based isolation and typing of *Shigella* using commercially available reagents has been the historic gold standard. New typing strategies include a qPCR-based approach directly either on cultured, single *Shigella* isolates or on stool samples, the latter of which provides advantages such as batched and centralized processing. This approach targets the *gtr* and *oac* O-antigen modification genes for serotyping *S. flexneri* (15–17), and holds several advantages over traditional microbiology including increased sensitivity of *Shigella* detection, batched and centralized sample processing, and reduced variability of culture due to differential abilities and laboratory capacities.

This study utilized data from the Enterics for Global Health (EFGH) site of Peru that concurrently serotyped *Shigella* using agglutination, a qPCR typing strategy on both cultured, single isolates and stool samples, and whole genome sequencing (WGS). Sequence data was analyzed using publicly available *in silico* tools and expert analysis to compare the performance of each method. Typing schemes were compared based on their ability to correctly characterize *Shigella* isolate serotypes.

## Materials and Methods

### Sample Origin

*Shigella* isolates were obtained as part of the EFGH study of Peru. This is an observational cross-sectional study designed to determine the incidence of medically attended shigellosis in children six to 35 months of age from seven different sites located in resource limited communities (18, 19). The EFGH study of Peru was conducted in Iquitos, Peru, between 2022 and 2024. Details of the site and population have been described previously (20).

### Ethics Statement

The study was approved by Ethics Review Committee of Asociacion Benefica Prisma and the Institutional Review Board of the University of Virginia. Written informed consent to participate in the study was obtained from the parents or legal guardians of children. Participants consented for further use of biological specimens.

### Serotyping methods

#### i. Culture and Agglutination based Serotyping

The microbiology methods of the EFGH study have been detailed previously (21). Briefly, rectal swabs were obtained using two nylon flocked swabs and each individually placed in Cary Blair and modified BGS transport medium (mBGS). Samples were transported to the laboratory for processing within 16 hours of collection at 2-8°C. Rectal swabs were used to inoculate both MacConkey (MAC) and XLD agar plates and incubated at 37°C for 24 hours. Non-lactose fermenting colonies with morphological characteristics compatible with *Shigella* were picked from both plates and re-streaked in clean fresh plates before initiating the biochemical test battery. Colonies were inoculated into motility indole ornithine (MIO), Kligler’s iron sugar (KIA), lysine iron agar (LIA) and urea agar. Colonies with Shigella compatible biochemistry were serotyped by agglutination with Denka Seiken polyvalent and *S. flexneri* monovalent antisera.

#### ii. Isolate based qPCR Serotyping

DNA extraction was completed from *Shigella* isolates using the crude lysate method. Specifically, 10 Shigella colonies cultured in trypticase soy agar (TSA) of each isolate were suspended in 200uL of 1mM EDTA buffer. Samples were incubated at 95°C for 15 minutes, centrifuged at 5000X g for 10 minutes and the supernatant was transferred into a new 1.5mL microcentrifuge tube for storage until qPCR testing was performed for *Shigella* serotyping and speciation as previously described (PMID 40157382). Briefly, multiplex qPCR testing was performed using the Ag-Path-ID One Step RT-PCR Kit (Thermo Fisher, Waltham, MA) in an QuantStudio 7 real-time PCR instrument (Applied Biosystems, Waltham, MA) under the following cycling conditions: 45 °C for 20 minutes, 95 °C for 10 minutes and 40 cycles of 95 °C for 15 seconds and 60 °C for 1 minute. Gene targets are shown in **Table 1**(22).

**Table 1.** qPCR panels used in the current study and the corresponding *Shigella* species and *S. flexneri* serotypes. The colours indicate the fluorophore of the probes, blue for FAM, green for VIC, red for Texas Red, and purple for Cy5.

| ***Shigella* species or**<br>***S. flexneri***<br>**serotype** | **qPCR panel I** | **qPCR panel II** | **qPCR panel III** |
| --- | --- | --- | --- |
| *S. flexneri* 1a |  | *gtrI* |  |
| *S. flexneri* 1b | *oac* | *gtrI* |  |
| *S. flexneri* 1d | *gtrX* | *gtrI* |  |
| *S. flexneri* 2a | *gtrII* |  |  |
| *S. flexneri* 2b | *gtrII* *gtrX* |  |  |
| *S. flexneri* 3a | *oac* *gtrX* |  |  |
| *S. flexneri* 3b | *oac* |  |  |
| *S. flexneri* 4a |  | *gtrIV* |  |
| *S. flexneri* 4b | *oac* | *gtrIV* |  |
| *S. flexneri* 5a | *oac* |  | *gtrV* |
| *S. flexneri* 5b | *oac* *gtrX* |  | *gtrV* |
| *S. flexneri* 6 | *wzx6* |  |  |
| *S. flexneri* 7a |  | *gtrI* | *gtrIc* |
| *S. flexneri* X | *gtrX* |  |  |
| *S. sonnei* |  | *S. sonnei* |  |
| *Shigella* |  | *ipaH* | *ipaH3* |

Serotype assignment was performed based on the following algorithm based on the optimization of the original scheme through EFGH (23):

1. The Cq of the *S. flexneri* serotyping target or *S. sonnei* target must be within 7.5 Cq of the ipaH Cq.
2. If ≥2 targets are required to determine the serotype, the Cq difference between the targets must be ≤6 Cq.

As noted in **Table 1**, this algorithm can identify *Shigella* spp., *S. sonnei*, *S. flexneri*, and *S. flexneri* serotypes: 1a, 1b, 1d, 2a, 2b, 3a, 3b, 4a, 4b, 5a, 5b, 6, 7a, and X. It cannot, however, identify *S. flexneri* Y, which is characterized by the absence of O-antigen modifications.

#### iii. Stool based qPCR serotyping

A third FLOQSwab rectal swab was obtained and placed in a dry tube for molecular identification of *Shigella* spp. These methods have also been previously detailed (23). Briefly, total nucleic acid (TNA) was extracted from rectal swabs using the QIAamp Fast DNA Stool mini kit (Qiagen, Hilden, Germany) with a pretreatment that included bead beating and 95°C incubation. TNA is then eluted with 200 μL of elution buffer. External controls, including 10^6^ phocine herpes virus (PhHV) and 10^7^ MS2 bacteriophage were spiked into each sample during the initial lysis step to monitor the extraction and amplification efficiency. One extraction blank was included per batch of extraction to assess for contamination. Samples were run using a TaqMan Array card with 82 targets, including *Shigella* species, and *S. flexneri* serotyping targets in a QuantStudio 7 real-time PCR instrument (Applied Biosystems, Waltham, MA). The assay master mix was prepared using Ag-Path-ID 2X RT-PCR buffer and Ag-Path-ID enzyme mix (Thermo Fisher, Waltham, MA), in addition to the TNA template. Cycling conditions were as follows: 45°C for 20 minutes and 95°C for 10 minutes, followed by 40 cycles of 95°C for 15 seconds and 60°C for 1 minute. Serotype assignment for attributable *Shigella* detection (*ipaH* Cq < 29.5) was performed as described for the isolate based qPCR serotyping(19). Additionally, if multiple *S. flexneri* serotypes and/or *S. sonnei* are detected using the above criteria, the target(s) with the lower Cq determines the primary species present.

**Figure 1** shows the *S. flexneri* serotype O-antigens, its modifications, and the genes utilized in qPCR for serotyping.

**Figure 1.**
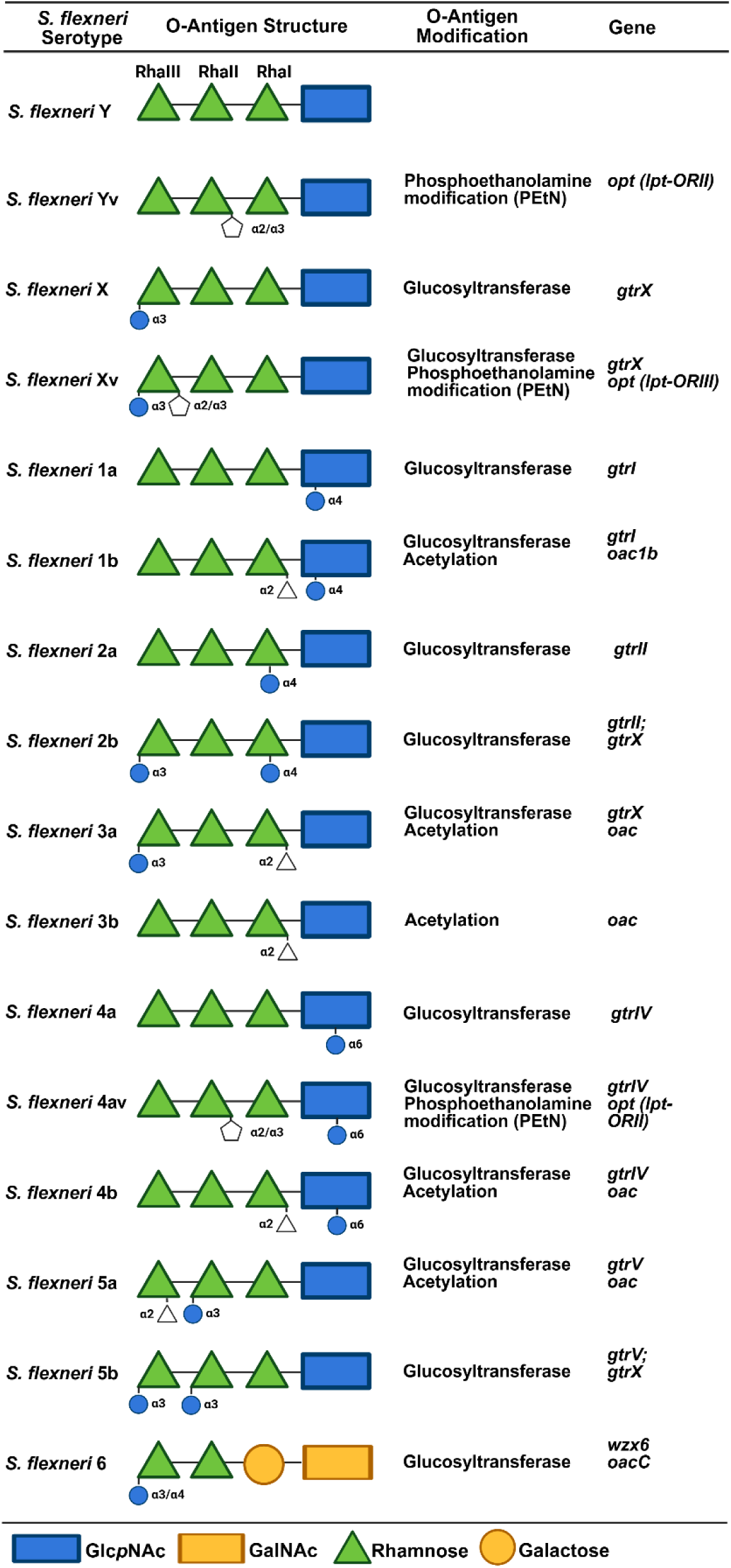
*Shigella flexneri* O antigen modifications and associated genes.

#### iv. Sequencing and in-silico genome-based prediction of serogroup and serotype

Genomic DNA from all isolates was purified using the Wizard genomic DNA purification kit according to the manufacturer’s directions (Promega, US). Genomic libraries were prepared using the Illumina DNA Prep Tagmentation kit as described previously (24) and sequenced using a MiSeq Reagent Kit v2 (500-cycles) on a MiSeq instrument (Illumina) at 16 pM, following the manufacturer’s protocols. Draft genomes are available at NCBI and are associated with BioProject PRJNA1321318 and accession numbers are in **Supplementary Table 1**

Reads were uploaded to Enterobase (http://enterobase.warwick.ac.uk/) (25) for assembly, quality control statistics and basic analysis. Additionally, assembled contigs were run through CheckM software v1.1.3 for completeness and contamination (26). Assembled contigs were run through ShigaPass v1.5.0, an in-silico tool to predict *Shigella* serotypes from whole genome assemblies (27).

#### v. Manual genome assessment serotype adjudication

Presence of *S. flexneri* O-antigen modification genes or mutations was performed using the Map to Reference command within Geneious Prime (v2025.1.2; Biomatters, Ltd., Auckland, New Zealand). Illumina paired-end reads >150 nt generated from an individual *S. flexneri* strain were mapped to 8 O-antigen biosynthesis and modification regions: *wba* (AE005674 (SF2096-SF2104)), *gtrABI* (CP130063 (QS169_18520-QS169_18530)), *oac1b* (JN377795), *gtrABII* (AE005674 (SF0305-SF0307)), *gtrX* (L05001), *oac* (AF547987), *gtrIV* (NC_022749 (V416_gp25)), and *opt* (KC020049). All *S. flexneri* serotypes 1–5, X and Y should have sequence reads that map to the *wba* locus, which encodes *S. flexneri* O-antigen backbone biosynthesis enzymes. Both *S. flexneri* 1a and *S. flexneri* 1b strains should have sequence reads that map to the *gtrABI* locus, and *S. flexneri* 1b should have sequence reads that map to *oac1b*. Both *S. flexneri* 2a and *S. flexneri* 2b strains should have sequence reads that map to the *gtrABII* and *S. flexneri* 2b should have sequence reads that map to *gtrX*. Both *S. flexneri* 3a and *S. flexneri* 3b strains should have sequence reads that map to *oac*, and *S. flexneri* 3a strains should have sequence reads that map to *gtrX*. *S. flexneri* 4a, *S. flexneri* 4av and *S. flexneri* 4b strains should have sequence reads that map to *gtrIV*, *S. flexneri* 4av strains should have sequence reads that map to *opt*, and *S. flexneri* 4b strains should have sequence reads that map to *oac*. *S. flexneri* Y strains have reads that only map to the *wba* locus and *S. flexneri* Yv strains should have sequence reads that map to *opt*. Genomic assemblies at 20X coverage were judged to be sufficient to reveal the absence/presence of O-antigen modification genes. Non-functional O-antigen modification genes due to premature STOP codons were identified with Geneious Prime at least 10X coverage across the entire gene of interest.

### Comparative Analysis

The identification of *S. flexneri* O-antigen modification genes and mutations performed in Geneious Prime (v2025.1.2; Biomatters, Ltd., Auckland, New Zealand) was considered the method to overrule discrepancies and definitive technique to adjudicate *Shigella* serogroups and serotypes. Agglutination, qPCR (on isolates and stool samples) and ShigaPass were compared to each other and ultimately to the gold standard.

A “*negative*” result is defined as the absence of the detection of *Shigella* in the specific sample analyzed. A “*not typed*” result is defined as a positive *Shigella* sample for which the test could not assign a serotype. The accuracy of each typing method was calculated as the total number of matching serotypes over the total number of samples evaluated by both compared methods, with 95% confidence intervals (CI) estimated using the Wilson score method. Additionally, the accuracy of each of the four serotyping methods for the *Shigella* serogroups and serotypes contained in the three main candidate vaccines (<u>candidate A</u>: *S. sonnei, S. flexneri* 1b, 2a and 3a (2); <u>candidate B</u>: *S. sonnei* and *S. flexneri* 2a. (3); <u>candidate C</u>: *S. sonnei*, *S. flexneri* 2a, 3a and 6 (4)) were estimated. Visualization and statistical analysis were performed in RStudio 2024.04.2.

## Results

Among the 1117 participants enrolled in the Peru EFGH study, 228 had a *Shigella* attributable diarrhea (10.2% (114/1117) culture-based prevalence; 19.2% (214/1117) qPCR-based prevalence). For this analysis we excluded children without a *Shigella* isolate, however serotypes assigned by stool-based qPCR are presented in **Supplemental Table 2.** Results of all four serotyping methods, (i) isolate based agglutination, (ii) isolate based qPCR serotyping, (iii) stool-based qPCR serotyping, and (iv) *in silico* genome-based serotyping, were available on 107 out of the 114 *Shigella* isolates (**Supplementary Table 1**). Whole genome sequencing was successfully performed for 111 out of the 114 available isolates, and isolate based qPCR was performed on 110 of the 114 isolates. All four methods identified a similar number of isolates as *S. flexneri*: agglutination identified 81/114 (71.1%), isolate-based qPCR 77/110 (70.0%), stool-based qPCR 76/114 (66.7%) and the *in silico* method 78/111 (70.2%). All methods identified 33 (28.9%) isolates as *S. sonnei*, except the stool-based qPCR which identified 32. The detailed number of *S. flexneri* serotypes identified by each serotyping method can be visualized in **Table 2**.

**Table 2.** *Shigella* serogroup and Serotyping results by serotyping technique.

| **Shigella Serogroup and Serotype** | **Definitive Serotype** | **Agglutination (n (%))** | **qPCR of Isolate (n (%))** | **qPCR of Stool (n (%))** | **In-silico (n (%))** |
| --- | --- | --- | --- | --- | --- |
| *S. flexneri* | 78 (68.4%) | 81 (71.1%) | 77 (67.5%) | 76 (66.7%) | 78 (68.4%) |
| 1a | 9 (11.5%) | 9 (11.1%) | 3 (3.9%) | 3 (3.9%) | 4 (5.1%) |
| 1b | 4 (5.1%) | 4 (4.9%) | 9 (11.7%) | 8 (10.5%) | 9 (11.5%) |
| 2a | 36 (46.2%) | 39 (48.1%) | 36 (46.8%) | 35 (46.1%) | 36 (46.2%) |
| 2b | 12 (15.4%) | 11 (13.6%) | 12 (15.6%) | 11 (14.5%) | 12 (15.4%) |
| 3a | 5 (6.4%) | 5 (6.2%) | 5 (6.5%) | 6 (7.9%) | 6 (7.7%) |
| 4a | 2 (2.6%) | 11 (13.6%) | 2 (2.6%) | 1 (1.3%) | 0 (0%) |
| 4av | 0 (0%) | 0 (0%) | 0 (0%) | 0 (0%) | 2 (2.6%) |
| 4b | 0 (0%) | 0 (0%) | 0 (0%) | 3 (3.9%) | 0 (0%) |
| Y | 3 (3.8%) | 2 (2.5%) | 0 (0%) | 0 (0%) | 2 (2.6%) |
| Yv | 7 (9.0%) | 0 (0%) | 0 (0%) | 0 (0%) | 7 (9.0%) |
| Not Typed | 0 (0%) | 0 (0%) | 10 (13.0%) | 9 (11.8%) | 0 (0%) |
| *S. sonnei* | 33 (28.9%) | 33 (28.9%) | 33 (28.9%) | 32 (28.1%) | 33 (28.9%) |
| Negative | 0 (0%) | 0 (0%) | 0 (0%) | 6 (5.3%) | 0 (0%) |
| Not Performed | 3 (2.6%) | 0 (0%) | 4 (3.5%) | 0 (0%) | 4 (2.6%) |
| **Total** | **114** | **114** | **114** | **114** | **114** |

Across all serotyping methods, the main misclassifications were associated with serotype Y (n=3) and serotype Yv (n=7). Specifically, agglutination-based serotyping classified all 7 Yv isolates as 4a and one of the three Y isolates also as 4a. The *in silico* based prediction tool correctly classified all 7 Yv isolates and 2 of the 3 Y isolates (one was incorrectly classified as a 1a). qPCR does not have the ability to serotype Y or Yv isolates. Thus, using isolate-based qPCR a total of nine Yv/Y isolates were “*non-typed*”, while one Y isolate was incorrectly classified as 1a. Similarly, stool-based qPCR serotyping provided a “*non-typed*” result for 5 Yv isolates and 2 Y isolates and incorrectly misclassified 2 Yv isolates as 4b and 1 Y isolate as 1a.

The second most common misclassification was associated with *S. flexneri* 1a. There were 9 isolates confirmed as 1a which were correctly classified as such by agglutination. However, the other three typing methods misclassified up to 7 of these as 1b. Manual evaluation of the contigs identified that the *oac* gene was present yet it contained a nonsense mutation. In-silico based serotype prediction misclassified three additional isolates: two *S. flexneri* 4a isolates were classified as 4av, yet manual examination of the contigs revealed the presence of the Petn-transferase moiety with a nonsense mutation, and one 1b isolate was classified as a 3a isolate, yet the *gtrI* and *oac* genes were confirmed to be present.

In rare instances, stool-based qPCR could not detect *Shigella* in stool, presumably due to stool sampling heterogeneity. Specifically, six samples were negative for the *ipaH* gene, which did not allow for stool-based genotyping. Additionally, in two instances the required cycle threshold was not achieved to call a serotype, obtaining a “*not typed’* output on a 1a isolate and on a 2a isolate.

Besides *S. sonnei*, *S. flexneri* 2a and 2b showed the highest concordance across all four serotyping methods. These two serotypes were also the most common *S. flexneri* serotypes in the current data set (2a: 46.2% (36/114); 2b: 15.4% (12/114)). Discrepancies associated with these two serotypes were limited.

**Figure 2** compares the results from the four serotyping methods to the gold standard of adjudicated manual sequence analysis. The accuracy of agglutination was 91.9% (102/111; 95% CI: 85.3% - 97.7%, Wilson), and that of ShigaPass was 91.0% (101/111; 95% CI: 84.2% - 95.0%, Wilson). The accuracy of the qPCR method on isolates was 91.8% (90/98; 95% CI: 84.7%-95.8%, Wilson). This calculation excludes excluding the *S. flexneri* Y and Yv samples (which carry no typable genes) that were accurately “*not-typed”* by the isolate based qPCR (n=9) When including the “*not-typed*” as discrepancies the accuracy was 84.1% (90/107; 95% CI: 76.0%-89.8%, Wilson). Similarly, the accuracy of the stool-based qPCR was 87.9% (87/99; 95% CI: 80.0%-92.9%, Wilson), when excluding “*negative*” (n=5) results and “*non-typed*” *S. flexneri* Y and Yv (n=7) from the analysis. Including these samples decreased accuracy to 78.3% (87/111; 95% CI: 69.8%-85.0%, Wilson). Complete concordance was achieved in 89.2% (83/93) of samples evaluated when excluding non-typed *S. flexneri* Y and Yv samples and negative outputs from qPCR (n=14), and in 77.6% (83/107) when including them. **Table 3** lists discrepancies and indicates the correct serotype assignment after identification of genes and/ or mutations causing discrepancies.

**Figure 2.**
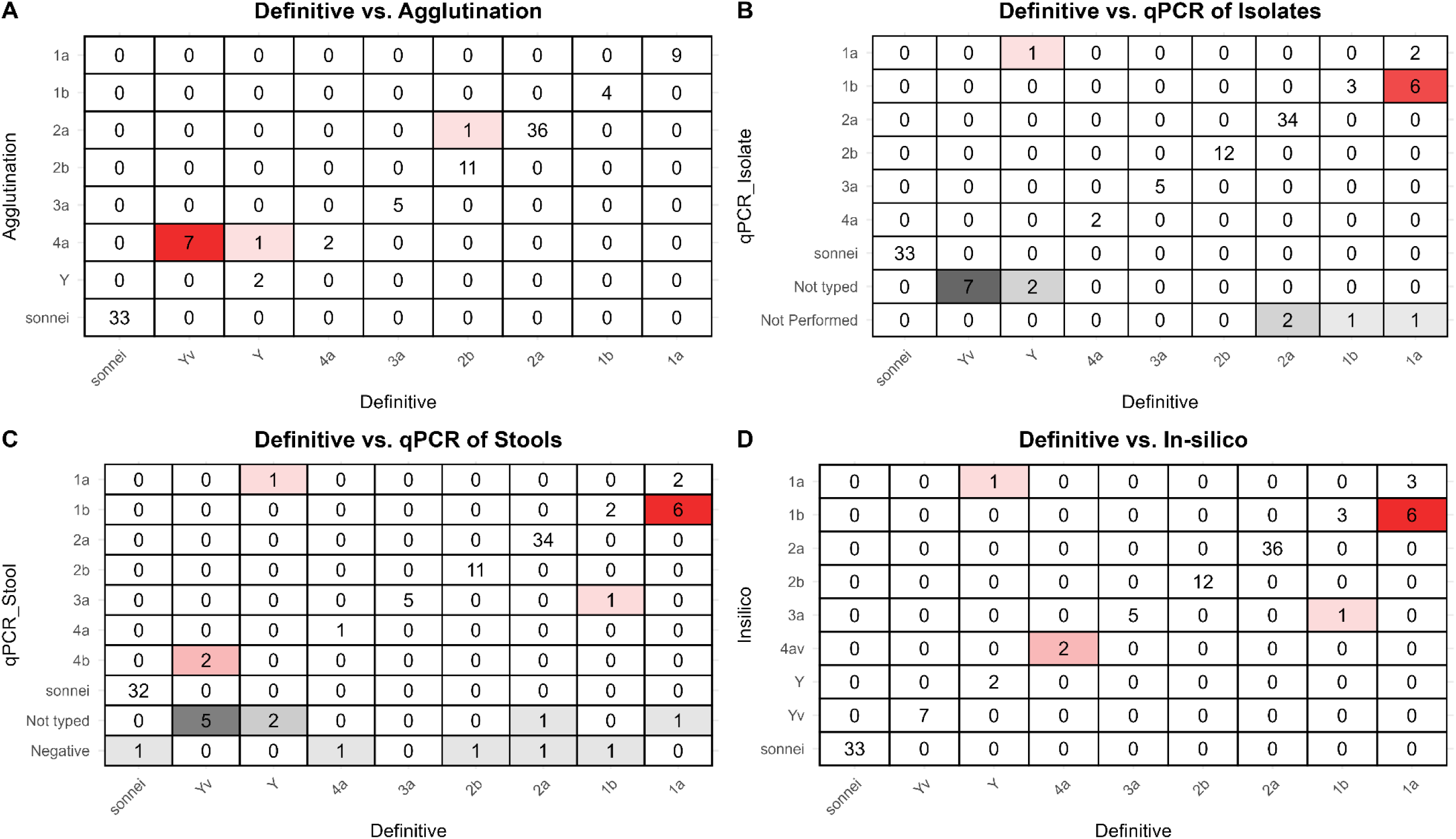
Serotype assignment of agglutination, isolate-based qPCR, stool-based qPCR and in-Silico in comparison to the definitive serotype adjudicated through manual sequence analysis. **Legend:** Comparison of the four serotyping tools used to define *Shigella* burden by serotype in disease burden studies and vaccine trials reveals that both agglutination and the in-silico tool assign correctly the same proportion of samples. All methods achieve >90% accuracy for Shigella serogroups and serotypes included in vaccine candidates.

**Table 3.** *Shigella* serotyping discrepancies between culture agglutination, qPCR and in-silico typing, and detection of genes and mutations leading to discrepancies.

| ID | Serogroup | Agglutination | qPCR of Isolate | qPCR of Stool | In-Silico | Definitive Serotype | Correct Assignment Explanation | Platform(s) with Correct Assignment |
| --- | --- | --- | --- | --- | --- | --- | --- | --- |
| **Discrepancies due to at least one serotyping mismatch** |  |  |  |  |  |  |  |  |
| 005005 | flexneri | 1a | 1b | 1b | 1b | 1a | *oac* (1b) gene w/ nonsense mutation | Agglutination |
| 005035 | flexneri | 1a | 1b | 1b | 1b | 1a | *oac* (1b) gene w/ nonsense mutation | Agglutination |
| 007901 | flexneri | 1a | 1b | 1b | 1b | 1a | *oac* (1b) gene w/ nonsense mutation | Agglutination |
| 014771 | flexneri | 1a | 1b | 1b | 1b | 1a | *oac* (1b) gene w/ nonsense mutation | Agglutination |
| 014681 | flexneri | 1a | 1b | 1b | 1b | 1a | *oac* (1b) gene w/ nonsense mutation | Agglutination |
| 014741 | flexneri | 1a | 1b | 1b | 1b | 1a | *oac* (1b) gene w/ nonsense mutation | Agglutination |
| 012476 | flexneri | 1a | NA | Not typed | 1a | 1a | *gtrI* gene present | Agglutination/ In-silico |
| 012103 | flexneri | 1b | 1b | 3a | 3a | 1b | *gtrI* and *oac* gene present | Agglutination/ qPCR of Isolate/ In-silico |
| 014581 | flexneri | 2a | 2b | Negative | 2b | 2b | *oacD* gene with frameshift | qPCR / In-silico |
| 014625 | flexneri | 2a | 2a | Not typed | 2a | 2a | *gtrII* gene present | Agglutination/ qPCR of Isolate/ In-silico |
| 002187 | flexneri | 4a | Not typed | Not typed | Yv | Yv | No *gtr* gene. Petn-transferase moiety present | In-silico |
| 006825 | flexneri | 4a | Not typed | Not typed | Yv | Yv | No *gtr* gene. Petn-transferase moiety present | In-silico |
| 006961 | flexneri | 4a | Not typed | Not typed | Yv | Yv | No *gtr* gene. Petn-transferase moiety present | In-silico |
| 007841 | flexneri | 4a | 4a | 4a | 4av | 4a | *gtrIV* gene present. Petn-transferase moiety present with a nonsense mutation | Agglutination/ qPCR |
| 011149 | flexneri | 4a | Not typed | Not typed | Yv | Yv | No *gtr* gene. Petn-transferase moiety present | In-silico |
| 014589 | flexneri | 4a | 4a | Negative | 4av | 4a | Has Petn-transferase but *lptO* gene it has a nonsense mutation | Agglutination/ qPCR of Isolate |
| 014593 | flexneri | 4a | Not typed | 4b | Yv | Yv | No *gtr* gene. Petn-transferase moiety present | In-silico |
| 014601 | flexneri | 4a | Not typed | 4b | Yv | Yv | No *gtr* gene. Petn-transferase moiety present | In-silico |
| 014633 | flexneri | 4a | Not typed | Not typed | Yv | Yv | No *gtr* gene. Petn-transferase moiety present | In-silico |
| 014697 | flexneri | 4a | Not typed | Not typed | Y | Y | No *gtr* gene | In-silico |
| 011803 | flexneri | Y | 1a | 1a | 1a | Y | No *gtr* gene | Agglutination |
| 000840 | flexneri | 4a | Not typed | 4b | NA | NA | Not available |  |
| 001671 | flexneri | 2a | 2a | Negative | NA | NA | Not available |  |
| **Discrepancies due to negatives and non-typable in stool samples only** |  |  |  |  |  |  |  |  |
| 009922 | flexneri | 1b | 1b | Negative | 1b | 1b | *gtrI* and *oac* gene present | Agglutination/ qPCR of Isolate/ In-silico |
| 014749 | flexneri | 2a | 2a | Negative | 2a | 2a | *gtrII* gene present | Agglutination/ qPCR of Isolate/ In-silico |
| 008864 | flexneri | Y | Not typed | Not typed | Y | Y | No *gtr* gene | Agglutination/ In-silico |
| 014163 | sonnei | sonnei | sonnei | Negative | sonnei | sonnei | S. sonnei | Agglutination/ qPCR of Isolate/ In-silico |

NA = Not Available

Finally, the accuracy of all serotyping methods exceeded 90% for all three vaccine combinations. Agglutination, isolate-based qPCR and *in silico* methods demonstrated almost perfect agreement for the serotypes included in these products (**Table 4**).

**Table 4.** Accuracy of all four serotyping methods for Shigella serogroups and serotypes included in current Shigella licensed vaccine and major vaccine candidates.

| Serogroup and Serotype combination | Number of Samples Fully Evaluated | Accuracy (% (n/N); 95% CI, Wilson) |  |  |  |
| --- | --- | --- | --- | --- | --- |
|  |  | *Agglutination* | *Isolate-based qPCR* | *Stool-based qPCR* | *In-silico* |
| Candidate A (*S. sonnei*; *S. flexneri* 1b, 2a and 3a) | 75 | 100% (75/75); 95.0-100% | 100 (75/75); 95.0-100% | 93.3% (70/75); 85.3%-97.1% | 98.7 (74/75); 92.8%-99.8% |
| Candidate B (*S. sonnei*; *S. flexneri* 2a) | 67 | 100% (67/67); 94.6-100% | 100 (67/67); 94.6-100% | 95.6% (64/67); 87.6%-98.5% | 100 (67/67); 94.6-100% |
| Candidate C (*S. sonnei*; *S. flexneri* 2a, 3a and 6) | 39 | 100% (39/39); 91.0-100% | 100 (39/39); 91.0-100% | 94.9% (37/39); 83.1%-98.6% | 100 (39/39); 91.0-100% |

## Discussion

Commercial antisera for serogrouping (*Shigella dysenteriae, Shigella flexneri, Shigella boydii*, and *Shigella sonnei*) and then serotyping of the isolates into serotypes involves agglutination, a test that has minimal logistical requirements and can be done readily at basic regional laboratories (28, 29). However, there are limitations of agglutination-based serotyping, which is costly and time-consuming, as well as harder to standardize across different laboratories(29, 30). Additionally, as shown in the EFGH study, culture sensitivity sensitivity compared to qPCR is considerably lower, observed agglutination reactions are also somewhat subjective, and cross-reactivity between serotypes is also possible. Indeed, we observed that antisera against *S. flexneri* 4a reacted to *S. flexneri* Yv strains that did not possess the *gtrIV* encoding the 1,6 glucosyltransferase that creates the 4a epitope (13). More importantly, there is no international standard and the companies producing antisera often discontinue production or do not possess antisera to newly discovered serotypes. Such is the case of serotypes Yv, 4av and Xv. These serotypes originate by the modification of the O-antigen with phosphoethanolamine (PEtN), which is encoded by the plasmid borne gene *opt* (7, 12). The difference in these serotypes is the position on the rhamnose that is substituted with PEtN. In both serotype Yv and 4a, the modification of the O-antigen is given at position 3 of rhamnose chain (Rha^III^), predominantly. In serotype Xv (not observed in this study), the O-antigen is also modified with PEtN, but on Rha^II^ (7, 12, 31). The differentiated pattern in phosphorylation is attributed to a difference of 11 bases (and 7 associated amino acid changes) in the gene sequences of the lpt-O gene homolog that is found in serotypes Yv and 4av (*opt*) versus serotype Xv (*lpt*-O_RII_). The availability of antisera to correctly type and identify these serotypes (Reagensia AB, Sweden) is no longer marketed, causing an incomplete categorization and profiling of *Shigella* serotypes that appears to have no clear solution. In the current study, the misidentification of serotype Yv as 4a was the most common serotyping error identified. Additionally, inconsistent typing results using different commercial and reference antisera is a well-documented phenomenon(16). These shortcomings speak to the value of moving to qPCR or sequence-based methodologies as more reproducible and rigorous over time and place, as the same isolate will consistently be able to be fully typed over time independent of market forces.

A practical alternative to serotyping based on agglutination is either a qPCR-based typing strategies or *in silico* analysis of whole genome sequencing of culture derived isolates. Both qPCR-based and *in silico* analysis classify serotypes according to *rfb* genes cluster sequences, bacteriophage seroconverting genes encoding O-antigen glycosylation and/ or O-acetylation enzymes that modify the O-antigen chain at specific positions (6, 8, 9, 11, 13), and plasmid encoded modification (7, 12). qPCR genotyping strategies, such as the qPCR serotyping strategy used here, have the advantage of a quick, single step serotype identification and has been found by other groups to have improved accuracy relative to agglutination when done on isolates (32). For state laboratories or vaccine trials, this method is the most efficiently to scale, limit costs and time spent typing isolates, as well as limit the cost associated with whole genome sequencing and associated expert requirements for bioinformatic sequencing. In the present analysis, isolate based qPCR serotyping performed similarly to agglutination and whole genome sequencing. It Is limited however, by its inability to detect Y and Yv, which have a relative low prevalence in most settings. The current qPCR algorithm is not designed to detect the Shigella O-antigen structure without any structural modification, as well as the absence of the *opt* gene that codes for PEtN from the PCR mixture. Additionally, it failed in the detection of a nonsense mutation in the *oac* gene reverting a 1b into a 1a serotype. However, this tool has the advantage of being customizable and adaptable to new *Shigella* serotypes and could potentially be improved to overcome current misclassifications. Moreover, the stool-based qPCR serotyping method is advantageous because it can serotype *Shigella* straight from fecal samples without the need for culturing (23, 33). Additionally, it shows approximately two-fold higher sensitivity relative to culture which may provide more robust estimates of serotype distribution, particularly in remote settings where specimen transport and culture may by logistically limited (34). Although its accuracy may be marginally lower than that of the three other serotyping strategies, this limitation is offset by the increased sensitivity of qPCR to detect Shigella in stool compared with culture. This would allow completion of a vaccine efficacy trial with fewer participants.

The WGS *in silico* serotyping tool utilized by this study, ShigaPass, presented certain limitations which are not commonly recognized. First, ShigaPass requires significant sequence coverage to correctly identify and serotype the *Shigella* isolates. Specifically, a WGS assembly obtained from *S. flexneri* 1b at 19x coverage using SPAdes assembler (35) and a 500 nt contig cutoff was not identified as *Shigella* by ShigaPass since a sufficient amount of *ipaH* was not contained on any contig. Second, ShigaPass does not detect nonsense mutations that make genes non-functional, and thus revert the serotype being encoded. For instance, ShigaPass identified two samples as serotype 4av, failing to identify a nonsense mutation in the *opt* gene that encodes for PEtN that would revert it to serotypes to 4a. Additionally, ShigaPass also identified 7 samples as *S. flexneri* seroptype 1b, failing to identify a nonsense mutation in the *oac* gene, reverting it to serotype 1a. When described in the literature, a correct assignment of serotype among 4879 genomes was observed in 98.5% and was noted to perform particularly well among *Shigella boydii* and *Shigella dysentariae* serogroups (27). Other *in silico* typing tools exist (ShigaTyper (36) and ShigEiFinder (37)), but are generally considered less accurate than ShigaPass (27). The differences between those attained by Yassine *et al* and the current analysis could be a function of the way genomes were selected. Yassine *et al* selected as reference strains from as many serotypes as available from all serogroups. The current analysis tested samples selected from a study that sampled children between the ages of 6-36 months presenting with diarrhea in a LMIC setting like one that would be considered for a vaccine trial. The relative distribution therefore reflects real world performance. In the current analysis, no *Shigella dysenteriae* was seen in this population, nor was there any *S. boydii*. The outcome metrics are thus driven by the distribution of serotypes in the target population for vaccine development and driven nearly entirely by the performance of the typing schemes for *S. flexneri* serogroups (all strategies performed well with *S. sonnei*) (27).

WGS provides the advantage of further genomic analysis. As mentioned above, we demonstrated that WGS reads could be mapped to the O-antigen biosynthesis and modification genes identifying nonsense mutations that would lead to eliminate the function of a gene that was scored as present by ShigaPass. Moreover, additional analysis provided within EnteroBase such as cgMLST, SNP calls, and antimicrobial resistance (AMR) markers can improve the understanding of the epidemiology of shigellosis, including phylogenomic investigations and prevalence and expansion of AMR. Although sequencing and bioinformatic analyses may involve higher costs and demand greater technical expertise, integrating an *in silico* tool such as ShigaPass into a web-based platform like EnteroBase or PubMLST could help overcome these costs, particularly as the cost of whole-genome sequencing continues to decline. That said, the relative increased cost, processing time, and bioinformatics restrict the utility of the widespread implementation of WGS for typing.

Vaccines under development are currently largely multivalent (38). Calculating serotype specific protection, and possible cross protection among antigenically related serogroups to those contained by the vaccine require accurate serotyping strategies. Considering the current licensed vaccine as well as two candidates in advanced stages of development (2–4), performance of all serotyping techniques is considered high and could be considered adequate for evaluating serotype specific protection during clinical trials.

The present study is limited by the limited number of serotypes found in Loreto, Peru. Thus, serotyping performance other serotypes not found in this geographic area could not be determined. That said, the serotypes found in this population are representative of the most frequently identified in large scale epidemiologic studies (39).

In conclusion, *in silico* tools such as ShigaPass provide a valuable step for serotyping *S. flexneri* genomes when complemented by mapping of DNA sequence reads to the O-antigen biosynthesis and modification genes of interest. Together these steps helped correctly serotype *Shigella* isolates in the absence of commercial antisera that was unavailable to perform appropriate and complete culture agglutination of isolates. We also note that qPCR-based serotyping is a valuable strategy can be expanded to new serogroups with additional primer construction and can inform serotyping in large-scale studies that cannot include culture and subsequent whole genome sequence *Shigella* spp.

## Data Availability

All data produced in the present work are contained in the manuscript.

## Acknowledgement

We would like to thank Dr. Nancy Strockbine for inputs on the early analysis and development of this work.

## Financial support

Funding for this study was provided by the National Institutes of Health of the United States (RO1AI158576 and R21AI163801 to MNK and CTP, D43TW010913 to MNK and MPO; K43TW012298 to FS). The Gates Foundation to PP (INV-016650 and INV-131791) and EH (INV-028721 and INV-041730) provided additional support to the EFGH study.

## Notes

### Competing Interest Statement

The authors have declared no competing interest.

### Author Declarations

Ethics committee/IRB of Asociacion Benefica Prisma and the University of Virginia gave ethical approval for this work.

## References

1. Collaborators GBDDD. Global, regional, and national age-sex-specific burden of diarrhoeal diseases, their risk factors, and aetiologies, 1990-2021, for 204 countries and territories: a systematic analysis for the Global Burden of Disease Study 2021. Lancet Infect Dis. 2025;25(5):519–36.

2. Leroux-Roels I, Maes C, Mancini F, Jacobs B, Sarakinou E, Alhatemi A, et al. Safety and Immunogenicity of a 4-Component Generalized Modules for Membrane Antigens Shigella Vaccine in Healthy European Adults: Randomized, Phase 1/2 Study. J Infect Dis. 2024;230(4):e971–e84.

3. Mo Y, Fang W, Li H, Chen J, Hu X, Wang B, et al. Safety and Immunogenicity of a Shigella Bivalent Conjugate Vaccine (ZF0901) in 3-Month- to 5-Year-Old Children in China. Vaccines (Basel). 2021;10(1).

4. Kelly M, Janardhanan J, Wagh C, Verma S, Charles RC, Leung DT, et al. Development of a Shigella conjugate vaccine targeting Shigella flexneri 6 that is immunogenic and provides protection against virulent challenge. Vaccine. 2024;42(24):126263.

5. MacLennan CA, Grow S, Ma LF, Steele AD. The Shigella Vaccines Pipeline. Vaccines (Basel). 2022;10(9).

6. Allison GE, Verma NK. Serotype-converting bacteriophages and O-antigen modification in *Shigella flexneri*. Trends Microbiol. 2000;8(1):17–23.

7. Sun Q, Lan R, Wang J, Xia S, Wang Y, Wang Y, et al. Identification and characterization of a novel Shigella flexneri serotype Yv in China. PLoS One. 2013;8(7):e70238.

8. Sun Q, Lan R, Wang Y, Wang J, Wang Y, Li P, et al. Isolation and genomic characterization of SfI, a serotype-converting bacteriophage of *Shigella flexneri*. BMC Microbiol. 2013;13:39.

9. Verma NK, Brandt JM, Verma DJ, Lindberg AA. Molecular characterization of the O-acetyl transferase gene of converting bacteriophage SF6 that adds group antigen 6 to *Shigella flexneri*. Mol Microbiol. 1991;5(1):71–5.

10. Adhikari P, Allison G, Whittle B, Verma NK. Serotype 1a O-antigen modification: molecular characterization of the genes involved and their novel organization in the Shigella flexneri chromosome. J Bacteriol. 1999;181(15):4711–8.

11. Mavris M, Manning PA, Morona R. Mechanism of bacteriophage SfII-mediated serotype conversion in *Shigella flexneri*. Mol Microbiol. 1997;26(5):939–50.

12. Sun Q, Knirel YA, Lan R, Wang J, Senchenkova SN, Jin D, et al. A novel plasmid-encoded serotype conversion mechanism through addition of phosphoethanolamine to the O-antigen of *Shigella flexneri*. PLoS One. 2012;7(9):e46095.

13. Adams MM, Allison GE, Verma NK. Type IV O antigen modification genes in the genome of Shigella flexneri NCTC 8296. Microbiology (Reading). 2001;147(Pt 4):851–60.

14. Levine MM, Kotloff KL, Barry EM, Pasetti MF, Sztein MB. Clinical trials of Shigella vaccines: two steps forward and one step back on a long, hard road. Nat Rev Microbiol. 2007;5(7):540–53.

15. Liu J, Pholwat S, Zhang J, Taniuchi M, Haque R, Alam M, et al. Evaluation of Molecular Serotyping Assays for Shigella flexneri Directly on Stool Samples. J Clin Microbiol. 2021;59(2).

16. Brengi SP, Sun Q, Bolanos H, Duarte F, Jenkins C, Pichel M, et al. PCR-Based Method for Shigella flexneri Serotyping: International Multicenter Validation. J Clin Microbiol. 2019;57(4).

17. Sun Q, Lan R, Wang Y, Zhao A, Zhang S, Wang J, et al. Development of a multiplex PCR assay targeting O-antigen modification genes for molecular serotyping of Shigella flexneri. J Clin Microbiol. 2011;49(11):3766–70.

18. Atlas HE, Conteh B, Islam MT, Jere KC, Omore R, Sanogo D, et al. Diarrhea Case Surveillance in the Enterics for Global Health Shigella Surveillance Study: Epidemiologic Methods. Open Forum Infect Dis. 2024;11(Suppl 1):S6–S16.

19. Yousafzai MT, Cornick J, Yori PP, Hossain MJ, Keita AM, Hannah EA, et al. Incidence and antimicrobial resistance of Shigella-attributable diarrhea in young children: Results from the multi-country enterics for global health (EFGH) Shigella Surveillance study. 2025.

20. Manzanares Villanueva K, Pinedo Vasquez T, Penataro Yori P, Romaina Cacique L, Garcia Bardales PF, Shapiama Lopez WV, et al. The Enterics for Global Health (EFGH) Shigella Surveillance Study in Peru. Open Forum Infect Dis. 2024;11(Suppl 1):S121–S8.

21. Horne B, Badji H, Bhuiyan MTR, Romaina Cachique L, Cornick J, Hotwani A, et al. Microbiological Methods Used in the Enterics for Global Health Shigella Surveillance Study. Open Forum Infect Dis. 2024;11(Suppl 1):S25–S33.

22. Rogawski McQuade ET, Liu J, Mahfuz M, Havt A, Varghese T, Shrestha J, et al. Epidemiology of Shigella species and serotypes in children: a retrospective substudy of the MAL-ED observational birth cohort study. Lancet Microbe. 2025;6(6):101064.

23. Liu J, Garcia Bardales PF, Islam K, Jarju S, Juma J, Mhango C, et al. Shigella Detection and Molecular Serotyping With a Customized TaqMan Array Card in the Enterics for Global Health (EFGH): Shigella Surveillance Study. Open Forum Infect Dis. 2024;11(Suppl 1):S34–S40.

24. Schiaffino F, Parker CT, Paredes Olortegui M, Pascoe B, Manzanares Villanueva K, Garcia Bardales PF, et al. Genomic resistant determinants of multidrug-resistant *Campylobacter* spp. isolates in Peru. J Glob Antimicrob Resist. 2024;36:309–18.

25. Zhou Z, Alikhan NF, Mohamed K, Fan Y, Agama Study G, Achtman M. The EnteroBase user’s guide, with case studies on Salmonella transmissions, Yersinia pestis phylogeny, and Escherichia core genomic diversity. Genome Res. 2020;30(1):138–52.

26. Parks DH, Imelfort M, Skennerton CT, Hugenholtz P, Tyson GW. CheckM: assessing the quality of microbial genomes recovered from isolates, single cells, and metagenomes. Genome Research. 2015;25(7):1043–55.

27. Yassine I, Hansen EE, Lefevre S, Ruckly C, Carle I, Lejay-Collin M, et al. ShigaPass: an in silico tool predicting Shigella serotypes from whole-genome sequencing assemblies. Microb Genom. 2023;9(3).

28. Bopp CA, Ries AA, Wells J, editors. Laboratory methods for the diagnosis of epidemic dysentery and cholera1999.

29. Kaminski RW, Pavlinac PB, Platts-Mills JA, Rogawski McQuade ET, Hausdorff WP, Isbrucker RA, et al. WHO Workshop Report: Regulatory Science to Inform Clinical Pathways for Shigella Vaccines Intended for Use in Children in Low- and Middle-Income Countries. Vaccines [Internet]. 2025; 13(5):[439 p.].

30. Bhuiyan TR, Liu J, Juma J, Horne BA, Hotwani A, Badji H, et al. Optimizing Shigella isolation: A multi-site evaluation of laboratory culture methods for Shigella detection, speciation, and serotyping with different transport media and sample types in the Enterics for Global Health study. medRxiv. 2025:2025.09.02.25334652.

31. Knirel YA, Lan R, Senchenkova SN, Wang J, Shashkov AS, Wang Y, et al. O-antigen structure of Shigella flexneri serotype Yv and effect of the lpt-O gene variation on phosphoethanolamine modification of S. flexneri O-antigens. Glycobiology. 2013;23(4):475–85.

32. Impact of malnutrition on small intestinal repair after acute viral enteritis. Nutrition reviews. 1985;43(11):335–7.

33. Liu J, Gratz J, Amour C, Kibiki G, Becker S, Janaki L, et al. A laboratory-developed TaqMan Array Card for simultaneous detection of 19 enteropathogens. J Clin Microbiol. 2013;51(2):472–80.

34. Liu J, Platts-Mills JA, Juma J, Kabir F, Nkeze J, Okoi C, et al. Use of quantitative molecular diagnostic methods to identify causes of diarrhoea in children: a reanalysis of the GEMS case-control study. Lancet. 2016;388(10051):1291–301.

35. Bankevich A, Nurk S, Antipov D, Gurevich AA, Dvorkin M, Kulikov AS, et al. SPAdes: a new genome assembly algorithm and its applications to single-cell sequencing. J Comput Biol. 2012;19(5):455–77.

36. Wu Y, Lau HK, Lee T, Lau DK, Payne J. In Silico Serotyping Based on Whole-Genome Sequencing Improves the Accuracy of Shigella Identification. Appl Environ Microbiol. 2019;85(7).

37. Zhang X, Payne M, Nguyen T, Kaur S, Lan R. Cluster-specific gene markers enhance Shigella and enteroinvasive Escherichia coli in silico serotyping. Microb Genom. 2021;7(12).

38. Martin P, Alaimo C. The Ongoing Journey of a Shigella Bioconjugate Vaccine. Vaccines (Basel). 2022;10(2).

39. Livio S, Strockbine NA, Panchalingam S, Tennant SM, Barry EM, Marohn ME, et al. Shigella isolates from the global enteric multicenter study inform vaccine development. Clin Infect Dis. 2014;59(7):933–41.

