## Supplementary Tables for "Robust Performance of Culture, qPCR, and Genomic Approaches for Shigella Serotyping in a Pediatric Surveillance Cohort"

**Supplementary Table 1.** *Shigella* spp. isolates and serotype assignment.

| BioSample | Accession | Sample ID | Period | Serogroup | Culture | Isolate TAC | Stool TAC | ShigaPass | Definitive | Genome Completeness | Genome Contamination | MLST |
|---|---|---|---|---|---|---|---|---|---|---|---|---|
| SAMN51230371 | SRR35310157 | 014581 | 10-Aug-22 | flexneri | 2a | 2b | Negative | 2b | 2b | 99.39 | 1.1 | ST-245 |
| SAMN51230372 | SRR35310156 | 014585 | 12-Aug-22 | flexneri | 2a | 2a | 2a | 2a | 2a | 99.35 | 0.26 | ST-6200 |
| SAMN51230373 | SRR35310145 | 014589 | 2-Sep-22 | flexneri | 4a | 4a | Negative | 4av | 4a | 98.66 | 0.53 | ST-630 |
| SAMN51230374 | SRR35310134 | 014593 | 11-Oct-22 | flexneri | 4a | Not typed | 4b | Yv | Yv | 98.76 | 1.05 | ST-630 |
| SAMN51230375 | SRR35310119 | 000840 | 13-Oct-22 | flexneri | 4a | Not typed | 4b | NA | NA | | | |
| SAMN51230376 | SRR35310108 | 014601 | 13-Oct-22 | flexneri | 4a | Not typed | 4b | Yv | Yv | 99.30 | 0.57 | ST-630 |
| SAMN51230377 | SRR35310125 | 014605 | 17-Oct-22 | sonnei | sonnei | sonnei | sonnei | sonnei | sonnei | 99.60 | 0.58 | ST-152 |
| SAMN51230378 | SRR35310124 | 014609 | 21-Oct-22 | sonnei | sonnei | sonnei | sonnei | sonnei | sonnei | 99.57 | 0.11 | ST-152 |
| SAMN51230379 | SRR35310123 | 014613 | 25-Oct-22 | sonnei | sonnei | sonnei | sonnei | sonnei | sonnei | 98.36 | 1.15 | ST-152 |
| SAMN51230380 | SRR35310122 | 001625 | 9-Nov-22 | flexneri | 2a | 2a | 2a | NA | NA | | | |
| SAMN51230381 | SRR35310155 | 014621 | 8-Nov-22 | flexneri | 2a | 2a | 2a | 2a | 2a | 99.38 | 0.26 | ST-6200 |
| SAMN51230382 | SRR35310154 | 014625 | 10-Nov-22 | flexneri | 2a | 2a | Not typed | 2a | 2a | 99.41 | 0.27 | ST-6200 |
| SAMN51230383 | SRR35310153 | 001671 | 10-Nov-22 | flexneri | 2a | 2a | Negative | NA | NA | | | |
| SAMN51230384 | SRR35310152 | 001820 | 17-Nov-22 | sonnei | sonnei | sonnei | sonnei | sonnei | sonnei | 99.60 | 0.11 | ST-152 |
| SAMN51230385 | SRR35310151 | 002171 | 25-Nov-22 | sonnei | sonnei | sonnei | sonnei | sonnei | sonnei | 99.60 | 0.58 | ST-152 |
| SAMN51230386 | SRR35310150 | 002174 | 25-Nov-22 | sonnei | sonnei | sonnei | sonnei | sonnei | sonnei | 99.60 | 0.11 | ST-152 |
| SAMN51230387 | SRR35310149 | 014633 | 25-Nov-22 | flexneri | 4a | Not typed | Not typed | Yv | Yv | 98.39 | 1.18 | ST-630 |
| SAMN51230388 | SRR35310148 | 002182 | 28-Nov-22 | sonnei | sonnei | sonnei | sonnei | sonnei | sonnei | 99.60 | 0.58 | ST-152 |
| SAMN51230389 | SRR35310147 | 002187 | 28-Nov-22 | flexneri | 4a | Not typed | Not typed | Yv | Yv | 99.27 | 0.57 | ST-630 |
| SAMN51230391 | SRR35310144 | 002370 | 6-Dec-22 | flexneri | 2b | 2b | 2b | 2b | 2b | 99.20 | 0.94 | ST-245 |
| SAMN51230392 | SRR35310143 | 002478 | 14-Dec-22 | flexneri | 2a | 2a | 2a | 2a | 2a | 99.35 | 0.26 | ST-6200 |
| SAMN51230393 | SRR35310142 | 002579 | 21-Dec-22 | flexneri | 2a | 2a | 2a | 2a | 2a | 99.43 | 0.26 | ST-6200 |
| SAMN51230394 | SRR35310141 | 002624 | 23-Dec-22 | flexneri | 2a | 2a | 2a | 2a | 2a | 99.40 | 0.26 | ST-6200 |
| SAMN51230395 | SRR35310140 | 002857 | 9-Jan-23 | flexneri | 2b | 2b | 2b | 2b | 2b | 99.39 | 0.94 | ST-245 |
| SAMN51230396 | SRR35310139 | 014641 | 24-Jan-23 | flexneri | 2b | 2b | 2b | 2b | 2b | 99.39 | 0.94 | ST-245 |
| SAMN51230397 | SRR35310138 | 014645 | 1-Feb-23 | flexneri | 2a | 2a | 2a | 2a | 2a | 99.70 | 0.26 | ST-245 |
| SAMN51230398 | SRR35310137 | 014649 | 6-Feb-23 | flexneri | 2a | 2a | 2a | 2a | 2a | 99.70 | 0.26 | ST-245 |
| SAMN51230399 | SRR35310136 | 014657 | 6-Feb-23 | flexneri | 2b | 2b | 2b | 2b | 2b | 99.39 | 0.94 | ST-245 |
| SAMN51230400 | SRR35310135 | 014661 | 7-Feb-23 | flexneri | 2a | 2a | 2a | 2a | 2a | 99.48 | 0.57 | ST-245 |
| SAMN51230401 | SRR35310133 | 014665 | 8-Feb-23 | flexneri | 2a | 2a | 2a | 2a | 2a | 99.70 | 0.66 | ST-245 |
| SAMN51230402 | SRR35310132 | 014669 | 13-Feb-23 | flexneri | 2a | 2a | 2a | 2a | 2a | 99.70 | 0.26 | ST-245 |
| SAMN51230403 | SRR35310131 | 014673 | 13-Feb-23 | flexneri | 2a | 2a | 2a | 2a | 2a | 99.69 | 0.26 | ST-245 |
| SAMN51230404 | SRR35310130 | 014677 | 14-Feb-23 | flexneri | 2a | 2a | 2a | 2a | 2a | 99.20 | 0.26 | ST-245 |
| SAMN51230405 | SRR35310129 | 014681 | 16-Feb-23 | flexneri | 1a | 1b | 1b | 1b | 1a | 99.36 | 0.31 | ST-245 |

| | | | | | | | | | | | | |
|---|---|---|---|---|---|---|---|---|---|---|---|---|
| SAMN51230406 | SRR35310128 | 014685 | 22-Feb-23 | flexneri | 2b | 2b | 2b | 2b | 2b | 99.39 | 0.94 | ST-245 |
| SAMN51230407 | SRR35310127 | 014689 | 24-Feb-23 | flexneri | 2b | 2b | 2b | 2b | 2b | 99.39 | 0.94 | ST-245 |
| SAMN51230408 | SRR35310126 | 014693 | 27-Feb-23 | flexneri | 2b | 2b | 2b | 2b | 2b | 98.80 | 2.77 | ST-245 |
| SAMN51230409 | SRR35310121 | 014697 | 2-Mar-23 | flexneri | 4a | Not typed | Not typed | Y | Y | 99.18 | 1.08 | ST-630 |
| SAMN51230410 | SRR35310120 | 014701 | 2-Mar-23 | sonnei | sonnei | sonnei | sonnei | sonnei | sonnei | 99.49 | 0.17 | ST-152 |
| SAMN51230411 | SRR35310118 | 014705 | 2-Mar-23 | sonnei | sonnei | sonnei | sonnei | sonnei | sonnei | 99.29 | 0.11 | ST-152 |
| SAMN51230412 | SRR35310117 | 014709 | 16-Mar-23 | flexneri | 3a | 3a | 3a | 3a | 3a | 98.98 | 0.39 | ST-628 |
| SAMN51230413 | SRR35310116 | 005005 | 29-Mar-23 | flexneri | 1a | 1b | 1b | 1b | 1a | 99.20 | 1.03 | ST-245 |
| SAMN51230414 | SRR35310115 | 014717 | 31-Mar-23 | flexneri | 2a | 2a | 2a | 2a | 2a | 99.75 | 0.26 | ST-245 |
| SAMN51230415 | SRR35310114 | 014721 | 31-Mar-23 | flexneri | 2b | 2b | 2b | 2b | 2b | 99.39 | 1.01 | ST-245 |
| SAMN51230416 | SRR35310113 | 005035 | 5-Apr-23 | flexneri | 1a | 1b | 1b | 1b | 1a | 99.33 | 0.31 | ST-245 |
| SAMN51230417 | SRR35310112 | 014729 | 10-Apr-23 | flexneri | 2a | 2a | 2a | 2a | 2a | 99.73 | 0.87 | ST-6200 |
| SAMN51230418 | SRR35310111 | 014733 | 12-Apr-23 | sonnei | sonnei | sonnei | sonnei | sonnei | sonnei | 99.57 | 0.29 | ST-152 |
| SAMN51230419 | SRR35310110 | 006007 | 19-Apr-23 | sonnei | sonnei | sonnei | sonnei | sonnei | sonnei | 94.67 | 2.96 | ST-152 |
| SAMN51230420 | SRR35310109 | 014741 | 19-Apr-23 | flexneri | 1a | 1b | 1b | 1b | 1a | 99.39 | 0.31 | ST-245 |
| SAMN51230421 | SRR35310107 | 014745 | 20-Apr-23 | flexneri | 2b | 2b | 2b | 2b | 2b | 99.05 | 1.26 | ST-245 |
| SAMN51230422 | SRR35310106 | 014749 | 2-May-23 | flexneri | 2a | 2a | Negative | 2a | 2a | 99.39 | 0.26 | ST-6200 |
| SAMN51231836 | SRR35312069 | 006335 | 12-May-23 | sonnei | sonnei | sonnei | sonnei | sonnei | sonnei | 99.44 | 0.11 | ST-152 |
| SAMN51231837 | SRR35312068 | 006545 | 22-May-23 | flexneri | 1a | 1a | 1a | 1a | 1a | 99.07 | 0.21 | ST-1024 |
| SAMN51231838 | SRR35312057 | 006550 | 22-May-23 | sonnei | sonnei | sonnei | sonnei | sonnei | sonnei | 99.60 | 0.11 | ST-152 |
| SAMN51231839 | SRR35312030 | 006735 | 30-May-23 | sonnei | sonnei | sonnei | sonnei | sonnei | sonnei | 99.60 | 0.30 | ST-152 |
| SAMN51231840 | SRR35312051 | 006825 | 31-May-23 | flexneri | 4a | Not typed | Not typed | Yv | Yv | 99.11 | 5.87 | ST-630 |
| SAMN51231841 | SRR35312040 | 006865 | 31-May-23 | flexneri | 2b | 2b | 2b | 2b | 2b | 99.39 | 0.94 | ST-245 |
| SAMN51231842 | SRR35312013 | 006961 | 7-Jun-23 | flexneri | 4a | Not typed | Not typed | Yv | Yv | 99.30 | 0.86 | ST-630 |
| SAMN51231843 | SRR35312009 | 007375 | 4-Jul-23 | flexneri | 2a | 2a | 2a | 2a | 2a | 99.43 | 0.26 | ST-6200 |
| SAMN51231844 | SRR35312008 | 007379 | 4-Jul-23 | sonnei | sonnei | sonnei | sonnei | sonnei | sonnei | 99.44 | 0.69 | ST-152 |
| SAMN51231845 | SRR35312007 | 007591 | 4-Jul-23 | flexneri | 2b | 2b | 2b | 2b | 2b | 99.39 | 1.05 | ST-245 |
| SAMN51231846 | SRR35312067 | 007841 | 25-Jul-23 | flexneri | 4a | 4a | 4a | 4av | 4a | 98.95 | 0.58 | ST-630 |
| SAMN51231847 | SRR35312066 | 007879 | 24-Jul-23 | flexneri | 2a | 2a | 2a | 2a | 2a | 99.70 | 1.19 | ST-245 |
| SAMN51231848 | SRR35312065 | 007883 | 31-Jul-23 | flexneri | 2a | 2a | 2a | 2a | 2a | 99.39 | 0.68 | ST-6200 |
| SAMN51231849 | SRR35312064 | 007901 | 24-Jul-23 | flexneri | 1a | 1b | 1b | 1b | 1a | 99.39 | 0.41 | ST-245 |
| SAMN51231850 | SRR35312063 | 014771 | 1-Aug-23 | flexneri | 1a | 1b | 1b | 1b | 1a | 99.32 | 0.67 | ST-245 |
| SAMN51231851 | SRR35312062 | 014775 | 15-Aug-23 | sonnei | sonnei | sonnei | sonnei | sonnei | sonnei | 99.60 | 0.11 | ST-152 |
| SAMN51231852 | SRR35312061 | 014779 | 17-Aug-23 | sonnei | sonnei | sonnei | sonnei | sonnei | sonnei | 99.53 | 0.32 | ST-152 |
| SAMN51231853 | SRR35312060 | 014783 | 24-Aug-23 | flexneri | 1a | 1a | 1a | 1a | 1a | 99.23 | 0.29 | ST-1024 |
| SAMN51231854 | SRR35312059 | 014787 | 24-Aug-23 | flexneri | 2a | 2a | 2a | 2a | 2a | 99.59 | 0.26 | ST-245 |
| SAMN51231855 | SRR35312058 | 014791 | 6-Sep-23 | flexneri | 2a | 2a | 2a | 2a | 2a | 99.70 | 0.26 | ST-6200 |

| | | | | | | | | | | | | |
|---|---|---|---|---|---|---|---|---|---|---|---|---|
| SAMN51231856 | SRR35312056 | 008524 | 8-Sep-23 | sonnei | sonnei | sonnei | sonnei | sonnei | sonnei | 99.60 | 0.11 | ST-152 |
| SAMN51231857 | SRR35312055 | 014795 | 19-Sep-23 | flexneri | 2a | 2a | 2a | 2a | 2a | 99.00 | 0.36 | ST-6200 |
| SAMN51231858 | SRR35312054 | 008864 | 26-Sep-23 | flexneri | Y | Not typed | Not typed | Y | Y | 99.27 | 0.00 | ST-630 |
| SAMN51231859 | SRR35312037 | 014803 | 26-Sep-23 | flexneri | 2a | 2a | 2a | 2a | 2a | 99.75 | 0.28 | ST-6200 |
| SAMN51231860 | SRR35312036 | 009076 | 6-Oct-23 | flexneri | 2a | 2a | 2a | 2a | 2a | 99.43 | 0.31 | ST-6200 |
| SAMN51231861 | SRR35312035 | 009191 | 13-Oct-23 | sonnei | sonnei | sonnei | sonnei | sonnei | sonnei | 99.60 | 0.11 | ST-152 |
| SAMN51231862 | SRR35312034 | 014807 | 16-Oct-23 | sonnei | sonnei | sonnei | sonnei | sonnei | sonnei | 99.60 | 0.58 | ST-152 |
| SAMN51231863 | SRR35312033 | 009554 | 30-Oct-23 | flexneri | 3a | 3a | 3a | 3a | 3a | 99.30 | 0.32 | ST-628 |
| SAMN51231864 | SRR35312032 | 009922 | 21-Nov-23 | flexneri | 1b | 1b | Negative | 1b | 1b | 99.39 | 0.31 | ST-245 |
| SAMN51231865 | SRR35312031 | 009955 | 22-Nov-23 | flexneri | 2a | 2a | 2a | 2a | 2a | 99.70 | 0.26 | ST-6200 |
| SAMN51231866 | SRR35312029 | 010178 | 1-Dec-23 | sonnei | sonnei | sonnei | sonnei | sonnei | sonnei | 99.60 | 0.11 | ST-152 |
| SAMN51231867 | SRR35312028 | 010204 | 4-Dec-23 | flexneri | 2a | NA | 2a | 2a | 2a | 99.70 | 0.29 | ST-17070 |
| SAMN51231868 | SRR35312027 | 010273 | 7-Dec-23 | sonnei | sonnei | sonnei | sonnei | sonnei | sonnei | 99.60 | 0.23 | ST-152 |
| SAMN51231869 | SRR35312026 | 010449 | 18-Dec-23 | sonnei | sonnei | sonnei | sonnei | sonnei | sonnei | 99.60 | 0.26 | ST-152 |
| SAMN51231870 | SRR35312025 | 010586 | 28-Dec-23 | flexneri | 3a | 3a | 3a | 3a | 3a | 99.30 | 0.36 | ST-628 |
| SAMN51231871 | SRR35312024 | 011018 | 16-Jan-24 | flexneri | 2a | 2a | 2a | 2a | 2a | 99.39 | 0.61 | ST-6200 |
| SAMN51231872 | SRR35312023 | 011022 | 16-Jan-24 | sonnei | sonnei | sonnei | sonnei | sonnei | sonnei | 99.60 | 0.14 | ST-152 |
| SAMN51231873 | SRR35312022 | 011141 | 18-Jan-24 | sonnei | sonnei | sonnei | sonnei | sonnei | sonnei | 99.24 | 0.11 | ST-152 |
| SAMN51231874 | SRR35312053 | 011149 | 18-Jan-24 | flexneri | 4a | Not typed | Not typed | Yv | Yv | 99.30 | 0.66 | ST-630 |
| SAMN51231875 | SRR35312052 | 011206 | 19-Jan-24 | flexneri | 2a | 2a | 2a | 2a | 2a | 99.39 | 0.26 | ST-6200 |
| SAMN51231876 | SRR35312050 | 011269 | 23-Jan-24 | flexneri | 2a | 2a | 2a | 2a | 2a | 99.70 | 0.29 | ST-6200 |
| SAMN51231877 | SRR35312049 | 011633 | 12-Feb-24 | flexneri | 1b | NA | 1b | 1b | 1b | 99.43 | 0.26 | ST-245 |
| SAMN51231878 | SRR35312048 | 011799 | 21-Feb-24 | flexneri | 2a | 2a | 2a | 2a | 2a | 99.39 | 0.26 | ST-6200 |
| SAMN51231879 | SRR35312047 | 011803 | 21-Feb-24 | flexneri | Y | 1a | 1a | 1a | Y | 99.39 | 0.31 | ST-245 |
| SAMN51231880 | SRR35312046 | 011807 | 21-Feb-24 | flexneri | 2a | 2a | 2a | 2a | 2a | 99.39 | 0.26 | ST-6200 |
| SAMN51231881 | SRR35312045 | 012095 | 27-Feb-24 | flexneri | 3a | 3a | 3a | 3a | 3a | 99.41 | 0.27 | ST-245 |
| SAMN51231882 | SRR35312044 | 012103 | 27-Feb-24 | flexneri | 1b | 1b | 3a | 3a | 1b | 99.41 | 0.27 | ST-245 |
| SAMN51231883 | SRR35312043 | 012111 | 27-Feb-24 | flexneri | 2a | 2a | 2a | 2a | 2a | 99.11 | 0.51 | ST-6200 |
| SAMN51231884 | SRR35312042 | 012298 | 20-Mar-24 | flexneri | 1b | 1b | 1b | 1b | 1b | 99.07 | 0.37 | ST-245 |
| SAMN51231885 | SRR35312041 | 012460 | 25-Mar-24 | flexneri | 2a | 2a | 2a | 2a | 2a | 99.43 | 0.29 | ST-6200 |
| SAMN51231886 | SRR35312039 | 012464 | 27-Mar-24 | flexneri | 3a | 3a | 3a | 3a | 3a | 99.33 | 0.32 | ST-628 |
| SAMN51231887 | SRR35312038 | 012468 | 2-Apr-24 | flexneri | 2a | 2a | 2a | 2a | 2a | 99.70 | 0.26 | ST-6200 |
| SAMN51231888 | SRR35312021 | 012476 | 4-Apr-24 | flexneri | 1a | NA | Not typed | 1a | 1a | 99.39 | 0.31 | ST-245 |
| SAMN51231889 | SRR35312020 | 012751 | 15-Apr-24 | flexneri | 2a | NA | 2a | 2a | 2a | 99.35 | 0.44 | ST-6200 |
| SAMN51231890 | SRR35312019 | 012895 | 24-Apr-24 | sonnei | sonnei | sonnei | sonnei | sonnei | sonnei | 99.60 | 0.43 | ST-152 |
| SAMN51231891 | SRR35312018 | 012958 | 2-May-24 | sonnei | sonnei | sonnei | sonnei | sonnei | sonnei | 99.49 | 0.58 | ST-152 |
| SAMN51231892 | SRR35312017 | 013047 | 8-May-24 | sonnei | sonnei | sonnei | sonnei | sonnei | sonnei | 99.60 | 0.29 | ST-152 |

| | | | | | | | | | | | | |
|---|---|---|---|---|---|---|---|---|---|---|---|---|
| SAMN51231893 | SRR35312016 | 013260 | 15-May-24 | sonnei | sonnei | sonnei | sonnei | sonnei | sonnei | 99.60 | 0.11 | ST-152 |
| SAMN51231894 | SRR35312015 | 013341 | 18-May-24 | flexneri | 2a | 2a | 2a | 2a | 2a | 99.70 | 0.26 | ST-6200 |
| SAMN51231895 | SRR35312014 | 014163 | 15-Jul-24 | sonnei | sonnei | sonnei | Negative | sonnei | sonnei | 99.60 | 0.11 | ST-152 |
| SAMN51231896 | SRR35312012 | 014167 | 17-Jul-24 | sonnei | sonnei | sonnei | sonnei | sonnei | sonnei | 99.60 | 0.11 | ST-152 |
| SAMN51231897 | SRR35312011 | 014349 | 31-Jul-24 | sonnei | sonnei | sonnei | sonnei | sonnei | sonnei | 99.60 | 0.11 | ST-152 |
| SAMN51231898 | SRR35312010 | 014403 | 7-Aug-24 | sonnei | sonnei | sonnei | sonnei | sonnei | sonnei | 99.58 | 0.12 | ST-152 |

**Supplementary Table 2.** Stool based qPCR serotype assignment.

| PID | Serotype |
|---|---|
| 6100216 | S.sonnei |
| 6100225 | S.sonnei |
| 6100305 | S.flexneri 2a |
| 6100331 | Untyped |
| 6100393 | S.flexneri 2a |
| 6100474 | Untyped |
| 6100495 | S.sonnei |
| 6100572 | S.flexneri 1b |
| 6100585 | S.flexneri 2a |
| 6100609 | S.sonnei |
| 6100611 | S.sonnei |
| 6100653 | S.flexneri 2a |
| 6100712 | S.sonnei |
| 6100773 | S.flexneri 2a |
| 6100845 | S.sonnei |
| 6100850 | Untyped |
| 6100861 | S.sonnei |
| 6100971 | S.flexneri 2a |
| 6101001 | S.flexneri 1b |
| 6101064 | S.sonnei |
| 6200092 | S.flexneri 2a |
| 6200136 | S.flexneri 4b |
| 6200160 | S.flexneri 4b |
| 6200212 | S.sonnei |
| 6200235 | Untyped |
| 6200321 | S.flexneri 2a |
| 6200363 | Untyped |
| 6200434 | S.sonnei |
| 6200465 | Untyped |
| 6200490 | S.flexneri 2b |
| 6200532 | S.flexneri 2a |
| 6200711 | S.flexneri 2a |
| 6200905 | S.flexneri 3a |
| 6200910 | S.sonnei |
| 6200968 | S.flexneri 1b |

| | |
|---|---|
| 6201011 | S.sonnei |
| 6201027 | S.flexneri 2b |
| 6201075 | S.sonnei |
| 6201121 | S.flexneri 1a |
| 6201139 | S.flexneri 1a |
| 6201142 | S.sonnei |
| 6201188 | S.sonnei |
| 6201195 | Untyped |
| 6201214 | Untyped |
| 6201238 | Untyped |
| 6201277 | S.sonnei |
| 6201371 | S.flexneri 1b |
| 6201405 | S.flexneri 1b |
| 6201422 | S.sonnei |
| 6201446 | S.sonnei |
| 6201468 | S.flexneri 2a |
| 6201479 | S.flexneri 2a |
| 6201493 | S.flexneri 1a |
| 6201529 | S.flexneri 2a |
| 6201565 | Untyped |
| 6201635 | S.flexneri 2a |
| 6201699 | Untyped |
| 6201702 | Untyped |
| 6201725 | Untyped |
| 6201778 | Untyped |
| 6201949 | Untyped |
| 6201960 | S.sonnei |
| 6201972 | S.sonnei |
| 6201985 | S.sonnei |
| 6201997 | S.sonnei |
| 6202074 | S.sonnei |
| 6202095 | S.flexneri 2a |
| 6202182 | Untyped |
| 6202267 | Untyped |
| 6202564 | S.sonnei |
| 6202607 | S.sonnei |
| 6300154 | S.flexneri 4b |

| | |
|---|---|
| 6300219 | S.flexneri 4b |
| 6300237 | S.flexneri 2a |
| 6300255 | S.sonnei |
| 6300262 | S.sonnei |
| 6300336 | S.sonnei |
| 6300397 | S.flexneri 2a |
| 6300459 | Untyped |
| 6300471 | S.sonnei |
| 6300521 | Untyped |
| 6300633 | Untyped |
| 6300657 | Untyped |
| 6300738 | Untyped |
| 6300882 | Untyped |
| 6300894 | S.sonnei |
| 6300918 | S.flexneri 2b |
| 6301014 | S.flexneri 2a |
| 6301023 | S.flexneri 2b |
| 6301092 | S.flexneri 2a |
| 6301185 | S.flexneri 2b |
| 6301220 | S.flexneri 2b |
| 6301356 | S.flexneri 4a |
| 6301465 | S.flexneri 2a |
| 6301476 | Untyped |
| 6301567 | S.sonnei |
| 6301580 | S.sonnei |
| 6301766 | S.sonnei |
| 6301775 | S.sonnei |
| 6301782 | S.sonnei |
| 6301794 | S.sonnei |
| 6301802 | S.sonnei |
| 6301816 | S.sonnei |
| 6301840 | Untyped |
| 6301869 | S.flexneri 1b |
| 6301878 | Untyped |
| 6301884 | S.flexneri 2a |
| 6301922 | S.flexneri 2a |
| 6301987 | S.sonnei |

| | |
|---|---|
| 6302051 | S.flexneri 4a |
| 6302085 | S.flexneri 1b |
| 6302161 | S.sonnei |
| 6302177 | S.flexneri 2a |
| 6302189 | S.flexneri 2a |
| 6302192 | S.flexneri 2a |
| 6302200 | S.flexneri 2a |
| 6302217 | S.flexneri 2a |
| 6302221 | Untyped |
| 6302301 | S.flexneri 2a |
| 6302364 | Untyped |
| 6302402 | Untyped |
| 6302528 | S.sonnei |
| 6302543 | S.sonnei |
| 6302581 | S.sonnei |
| 6302698 | S.flexneri 3a |
| 6302710 | S.flexneri 2a |
| 6302722 | S.sonnei |
| 6302746 | Untyped |
| 6302852 | S.flexneri 1b |
| 6302890 | S.flexneri 1a |
| 6302909 | S.flexneri 2a |
| 6302953 | S.flexneri 3a |
| 6302966 | S.flexneri 3a |
| 6303025 | Untyped |
| 6303069 | S.flexneri 2a |
| 6303084 | S.flexneri 2a |
| 6303198 | Untyped |
| 6303318 | S.sonnei |
| 6303352 | S.flexneri 2a |
| 6303408 | S.flexneri 2a |
| 6303692 | S.sonnei |
| 6303728 | S.sonnei |
| 6400133 | S.sonnei |
| 6400169 | S.sonnei |
| 6400229 | Untyped |
| 6400330 | S.flexneri 1b |

| | |
|---|---|
| 6400366 | S.flexneri 1b |
| 6400401 | S.flexneri 2a |
| 6400538 | S.sonnei |
| 6400545 | S.sonnei |
| 6400646 | S.flexneri 2a |
| 6400654 | S.flexneri 1b |
| 6400668 | Untyped |
| 6400687 | S.flexneri 2a |
| 6400721 | S.sonnei |
| 6400742 | S.flexneri 2a |
| 6400808 | Untyped |
| 6400919 | S.flexneri 3a |
| 6401088 | S.sonnei |
| 6401194 | S.flexneri 2a |
| 6500027 | S.flexneri 2a |
| 6500142 | S.flexneri 4a |
| 6500174 | Untyped |
| 6500292 | S.flexneri 2a |
| 6500332 | S.sonnei |
| 6500380 | Untyped |
| 6500405 | S.sonnei |
| 6500410 | S.sonnei |
| 6500446 | Untyped |
| 6500493 | S.flexneri 2a |
| 6500583 | S.flexneri 2b |
| 6500702 | S.flexneri 2a |
| 6500716 | S.flexneri 2a |
| 6500740 | S.flexneri 2a |
| 6500769 | S.flexneri 2a |
| 6500784 | S.sonnei |
| 6500791 | S.flexneri 2a |
| 6500896 | S.flexneri 2b |
| 6500908 | S.flexneri 1b |
| 6500913 | S.sonnei |
| 6500985 | S.sonnei |
| 6501040 | S.sonnei |
| 6501057 | S.flexneri 1a |

| | |
|---|---|
| 6501078 | S.flexneri 2b |
| 6501084 | Untyped |
| 6501091 | S.flexneri 2b |
| 6501103 | Untyped |
| 6501115 | Untyped |
| 6501126 | Untyped |
| 6501159 | Untyped |
| 6501209 | S.flexneri 2b |
| 6501227 | Untyped |
| 6501318 | S.flexneri 1b |
| 6501341 | S.sonnei |
| 6501352 | S.flexneri 1a |
| 6501365 | S.flexneri 2a |
| 6501376 | S.flexneri 2a |
| 6501424 | Untyped |
| 6501522 | S.sonnei |
| 6501645 | S.sonnei |
| 6501704 | S.flexneri 3a |
| 6501728 | S.flexneri 2a |
| 6501743 | S.sonnei |
| 6501755 | S.sonnei |
| 6501762 | S.flexneri 2a |
| 6501770 | S.flexneri 2a |
| 6501858 | S.flexneri 1b |
| 6501886 | S.flexneri 2a |
| 6501939 | S.flexneri 2a |
| 6501988 | Untyped |
| 6502003 | S.flexneri 3a |
| 6502015 | S.flexneri 2a |
| 6502098 | S.sonnei |
| 6502290 | S.sonnei |
